# What role do adolescents’ independent food purchasing choices play in their dietary quality?

**DOI:** 10.1101/2024.10.17.24315666

**Authors:** Sarah Shaw, Sarah Crozier, Cyrus Cooper, Dianna Smith, Mary Barker, Christina Vogel

## Abstract

During adolescence, many young people start to make more independent food purchases. Subsequently, these independent food choices will increasingly contribute to their overall diet quality; little is known, however, about this relationship. This study aimed to examine the role adolescents’ independent food purchases play in their diet quality and assess if these relationships vary according to socio-economic status.

A convenience sample of adolescents aged 11-18 years and attending secondary school or college in Hampshire, England, were recruited to participate in a one-week cross-sectional observational study. A validated 20-item Food Frequency Questionnaire assessed diet quality. Participants used an ecological momentary assessment mobile phone app to record food purchases.

Over seven days, 552 food/drink items were purchased on 253 food purchasing occasions by 80 participants. The majority of purchases (n=329, 60%) were coded as ‘not adhering’ to the UK Eatwell Guide, 32% were coded as ‘adhering’ and 9% fell between these two categories. Healthier food purchasing was associated with better diet quality (β 0.48, (95%CI 0.01, 0.96) p=0.05); results attenuated after adjustment for confounders (β 0.36, (95%CI −0.15, 0.87) p=0.16). Interaction models showed purchasing healthfulness was more strongly associated with diet quality among young people of lower SES (p=0.01). The majority of adolescents’ independent food purchases did not adhere to healthy eating guidelines. For adolescents experiencing socioeconomic disadvantage, these food choices had a more detrimental impact on their overall diet quality than they did for more advantaged adolescents. Future research should focus on identifying ways to support more healthful independent food choices by adolescents to reduce dietary inequalities and improve health and well-being among the next generation of adults.

## Introduction

Poor dietary behaviours are a major contributor to the global burden presented by non-communicable diseases (NCDs) (1). Evidence from the UK’s National Diet and Nutrition Survey suggests that adolescents have poorer dietary behaviours when compared to other age groups (2, 3). The dietary habits of this age group are an important public health issue as behaviours established during this period frequently track into adulthood and play a role in future health trajectories (4). Identifying ways to support healthy dietary behaviours among adolescents is therefore an important strategy to improve health and reduce the prevalence of NCDs; intervening in adolescence has potential to improve the immediate and future health of the individual as well as the health of any future offspring (5).

Adolescence is a time when individuals gain autonomy and independence. During this stage of the lifecourse, many young people start making more of their own food choices without supervision from parents or other care givers. While it has been shown that adolescents source the majority of their food from home and school (6–8), many adolescent autonomous food decisions are likely to be made outside of these home and school environments. A 2015 report from Food Standards Scotland showed that the majority of independent food choices made by adolescents outside of the home and school at lunchtimes are high in fat, sugar and salt; chips (fries), sugary soft drinks and energy drinks were some of the most frequently purchased items (*9*). Independent food choices made by adolescents are likely to represent an increasing proportion of their overall food intake and, therefore, their overall diet quality. There is limited evidence, however, about how the foods adolescents purchase for themselves affect their overall dietary quality. Findings from a study in Kentucky, USA, showed that adolescents with unhealthy shopping habits (shopping three or more times per week at petrol stations, convenience stores and fast-food restaurants) ate fewer fruits and vegetables than those with healthier shopping habits (10). Understanding the contribution that independent food purchases of adolescents make to their overall dietary intake is important to design interventions and food policies that are effective at improving the diet and health of this important age group.

Socio-economic status (SES) has been well documented as a critical determinant of dietary quality. In the UK, adolescents living in more deprived areas and lower income households have been shown to have poorer quality diets (11–13), in particular consuming lower levels of vegetables, fibre and oily fish and more energy drinks (11–13). However, the role that adolescents’ independent food purchasing decisions play in their overall diet quality and the difference by SES remains unclear.

In order to address this knowledge gap, this study aimed to:

1. Examine food purchasing behaviours as predictors of diet quality;
2. Assess whether SES moderated the relationship between food purchasing behaviours and diet quality of adolescents.

## Material and Methods

### Study design

A one-week cross-sectional, observational study was conducted to collect data on adolescents’ independent food purchasing behaviours and diet quality as part of the Map My Food Study. Ethical approval for this study was granted by the Faculty of Medicine Ethics Committee, University of Southampton (ERGO 57044).

### Participants and recruitment

A convenience sample of adolescent participants was recruited from schools, colleges and youth groups based in Hampshire in the south of England, UK. Adolescents were eligible to participate if they i) were aged 11-18 years, ii) attended school or college in England and iii) owned a GPS-enabled smart phone.

A researcher attended the school, college or youth group to introduce the study and answer questions. Information was provided about the purpose and methods of the study. Written informed consent was obtained for all participants using an electronic consent form. Participants over the age of 16 years were able to provide consent to participate. Those aged under 16 years provided assent and parental consent was also sought for their participation.

### Data collection

Data were collected between May 2021 and April 2022. Following the consent process, participants were asked to complete an online questionnaire containing demographic questions and a food frequency questionnaire (FFQ). Participants were also asked to download and use a mobile phone app for one week. The app was a modified version of the Tripzoom app which was created and run by a commercial company, Locatienet (14, 15). Where possible, a member of the research team aimed to be present when the participants were completing the questionnaire and downloading the study app. On some occasions, restrictions on visitors entering schools and colleges during the COVID-19 pandemic prevented this happening. In these instances, participants were able to contact the research team via email if they experienced any difficulties.

The app used Ecological Momentary Assessment (EMA) methods to collect data about the food outlets adolescents used and their food purchases over the study week. EMA is an intensive longitudinal data collection technique aimed at reducing recall bias by collecting data from study participants when they are in their natural environment, and close in time to when the behaviour of interest takes place (16). EMA has been used previously with adolescents and young adults in order to provide a better understanding of the external and internal cues influencing dietary consumption (17–20). Adolescents were asked to use the app to record all their independent food and beverage purchases over the study week. Three time-based prompts were delivered through the app to remind participants to make a recording and to reduce incidences of missing data. To make a recording, adolescents could select a food or drink from a pre-defined list of food items. If an item was not present on the list, details could be entered using a free text field. Participants could also take photos of the items or receipts. These photos were used to check the accuracy of the recorded food items. In total, photos were provided for 23% of entries. With each entry, participants were also asked to provide details about the purpose of the purchase (breakfast, lunch, dinner or snack). The app data collection methods were refined through feasibility testing (n=14) and a pilot study (n=12) with young people to ensure acceptability (unpublished data). At the end of the study week, participants who had completed the questionnaire and downloaded the app were given a £10 Amazon voucher as an appreciation for their participation in the study.

### Exposure Measures

#### Demographic data

Using the questionnaire, participants provided their age, gender, ethnicity and home postcode. Household-level SES was assessed using questions from the Family Affluence Scale (FAS) (21). These questions have been used in the UK What about YOUth? Survey and the International Health Behaviour in School-aged Children study (21). The questions have been validated and are shown to be easier to complete than questions about parental education and occupation (21). A binary score was created to represent household SES: low household SES (FAS 1-7), high household SES (FAS 8-14). The Index of Multiple Deprivation (IMD) (22), the official measure of relative deprivation for small geographical areas in England, was calculated for each participant using their home postcode. A binary score was created to represent neighbourhood SES: low neighbourhood SES (IMD scores 1-5), high neighbourhood SES (IMD scores 6-10).

#### Total number of food purchases

Using the food purchasing data recorded in the app, the number of food/drink items purchased over the study week was tallied for each participant. First, the number of items purchased on each food purchasing occasion was tallied; for example, if a participant purchased a burger, chips, and a sugar sweetened beverage, three items would be recorded for that food purchasing occasion. These numbers were then tallied for all food purchasing occasions made by each participant to give the total number of food purchases made over the study week.

#### Healthfulness of food purchases

All food and drink items purchased during the Map My Food Study were classified according to how well they adhered to the UK’s Eatwell Guide (23). Food and drink items were classified as either ‘adhering’, ‘not-adhering’ or ‘combination’. Food and beverage items which are included as part of the main food groups in the Eatwell Guide and those that individuals are encouraged to eat more were categorised as ‘adhering’. Foods high in fat, sugar and salt that fall outside the main food groups in the Eatwell guide and which individuals are encouraged to limit were categorised as ‘not-adhering’. Foods that could not be classified within either of these groups were classified as ‘combination’ foods.

Table 1 shows the food purchases recorded by participants and how they were classified in terms of healthfulness. Previous research has used similar coding approaches based on the Australian Guide to Healthy Eating (6, 24).

**Table 1.** Coding of food purchases against the UK Eatwell Guide.

| Coding food purchases against Eatwell Guide |  |  |
| --- | --- | --- |
| Foods/drinks categorised as adhering to the Eatwell Guide | Foods/drinks categorised as not adhering to the Eatwell Guide | Combination/uncategorised foods/drinks |
| • Bread<br>• Breakfast cereal (No added sugar)<br>• Fermented milk drink<br>• Fruit<br>• Fruit Juice<br>• Fruit Smoothie<br>• Non-sugar sweetened carbonated drink<br>• Milk<br>• Nuts<br>• Pasta/rice<br>• Sushi<br>• Tea/Coffee<br>• Steak<br>• Roasted chicken<br>• Vegetables<br>• Water | • Battered Fish<br>• Biscuits<br>• Burgers/kebabs<br>• Cakes<br>• Chicken nuggets/battered chicken<br>• Chips/Fries<br>• Chocolate confectionary<br>• Crisps<br>• Energy drinks<br>• Ice-cream<br>• Iced Tea/ Iced Coffee/ Bubble Tea<br>• Milkshakes<br>• Non-carbonated sugar sweetened drinks<br>• Popcorn<br>• Pizza<br>• Processed meat (chorizo)<br>• Ready meals<br>• Sauces and dips<br>• Sugar confectionary<br>• Sausage roll, pasty, pie<br>• Sugar sweetened carbonated drinks<br>• Sweet pastries | • Sandwich or wrap<br>• Other hot drink<br>• Dumplings |

To create Purchasing Healthfulness Scores, foods classified as ‘adhering’ were allocated the score of ‘+1’, the ‘combination’ classification was allocated the rating of ‘0’ and the ‘non-adhering’ foods were allocated the rating of ‘-1’. Food scores were subsequently tallied for each food purchasing occasion and then divided by the total number of food/drink items purchased as part of that food purchasing occasion. The same process was used to calculate a Weekly Purchasing Healthfulness Score for each participant; food scores were tallied for the entire week and then divided by the total number of food/drink items purchased over the week. Scores ranged between −1 and 1 and were used as continuous variables and presented as healthfulness units. Higher scores representing healthier independent purchasing patterns.

### Outcome Measures

#### Diet quality

Using a 20-item FFQ, participants recorded how frequently they consumed each food group over the previous month. These dietary data were used to calculate a diet quality score for each participant following published methodology specifically developed to assess diet quality among UK adolescents (11). This diet quality score has been validated against fourteen nutritional biomarkers, including serum folate, homocysteine, total carotenoids and vitamins B12, C and D (11).

### Statistical analysis

Normally-distributed continuous variables are summarised using mean (SD). Non-normally distributed continuous variables are summarised using median (IQR).

Categorical variables are summarised using n (%). T-tests, Mann-Whitney rank sum tests and Chi-squared tests were used to compare differences in demographic characteristics between those with purchasing data and those without. Differences in Diet Quality Scores and purchasing outcomes according to demographic characteristics were assessed using unpaired t-tests and Mann-Whitney rank-sum tests. In order to retain statistical power, a test for trend was conducted for characteristics measured using an underlying continuous variable (age, IMD, and household SES).

To address aim 1, two separate linear regression models were fitted both using the Diet Quality Score as the outcome. The exposure for the first regression model was the variable describing the number of food purchases over the study week. The exposure for the second model was the Weekly Purchasing Healthfulness Score. Age, gender, ethnicity, and household SES were included as confounding variables in adjusted models, reflecting their observed influence in previous studies (11, 25).

To address aim 2, an interaction term for Weekly Purchasing Healthfulness Score by household SES was added to the regression models to determine the effect modification of household SES in the relationship between food purchasing behaviours and diet quality.

All data were analysed in Stata version 16 (26).

## Results

### Participant Characteristics

A total of 108 participants completed the online questionnaire. Of these, 101 participants downloaded the study app and food purchasing data were available for 80 participants. Table 2 shows the demographic characteristics for participants with and without purchasing data. The majority of participants with both purchasing and dietary data were girls (69%), self-identified as being of white ethnicity (79%) and had a median age of 17 years (IQR 15, 17). Thirty participants (37%) were living in homes located in the most deprived half of neighbourhoods in the UK. Twenty-four participants (30%) had a household SES (Family Affluence Score) between 0-7, representing low household-level SES.

**Table 2.** Characteristics of study participants.

| Characteristics | Participants with purchasing and diet data (n=80) | Participants without purchasing data (n=28) |
| --- | --- | --- |
| **Gender, n (%)** |  |  |
| Boy | 19 (24) | 14 (50) |
| Girl | 55 (69) | 13 (46) |
| Not provided | 6 (7) | 1 (3) |
| **Age, median (IQR) (years)** | 17 (15,17) | 14 (13,17) |
| **Age, n (%) (years)** |  |  |
| 11-13 years | 8 (10) | 9 (32) |
| 14-16 years | 25 (31) | 10 (36) |
| 17-18 years | 47 (59) | 9 (32) |
| **Ethnicity, n (%)** |  |  |
| White | 63 (79) | 25 (89) |
| Other ethnicities* | 17 (21) | 3 (11) |
| **Area-level SES (IMD), n (%)** |  |  |
| 1-5 (most deprived) | 30 (37) | 12 (43) |
| 6-10 (least deprived) | 34 (43) | 13 (46) |
| Missing | 16 (20) | 3 (11) |
| **Household-level SES (FAS), n (%)** |  |  |
| Low (0-7) | 24 (30) | 5 (18) |
| High (8-14) | 54 (68) | 23 (82) |
| Missing | 2 (2) | 0 (0) |

### Food purchasing behaviours

In total, 552 food/drink items were purchased during 253 food purchasing occasions over the study week. The median number of purchases made by each participant over the 7-day period was 5 (IQR 3, 8; range 1-30). As visualised in Fig 1, the most frequently purchased foods over the study period were chips (n= 47, 9% of all purchases), sandwiches (n=40, 7% of all purchases) and vegetables (n= 30, 5% of all purchases). The most frequently purchased drinks were non-sugar-sweetened carbonated drinks (n=28, 5% of all purchases), sugar-sweetened carbonated drinks (n=24, 4% of all purchases) and water (n=19, 3% of all purchases). The majority of purchases (n= 329, 60%) were coded as ‘not adhering’ to the UK Eatwell Guide, 175 food items (32%) were coded as ‘adhering’ and 48 (9%) were coded as combination foods. Fig 2 presents the purchases made by participants of high and low household SES, by the three UK Eatwell classifications. This graph shows no difference in percentage of purchased foods categorised as adhering, not adhering and combination between participants with high and low household SES.

**Fig 1.**
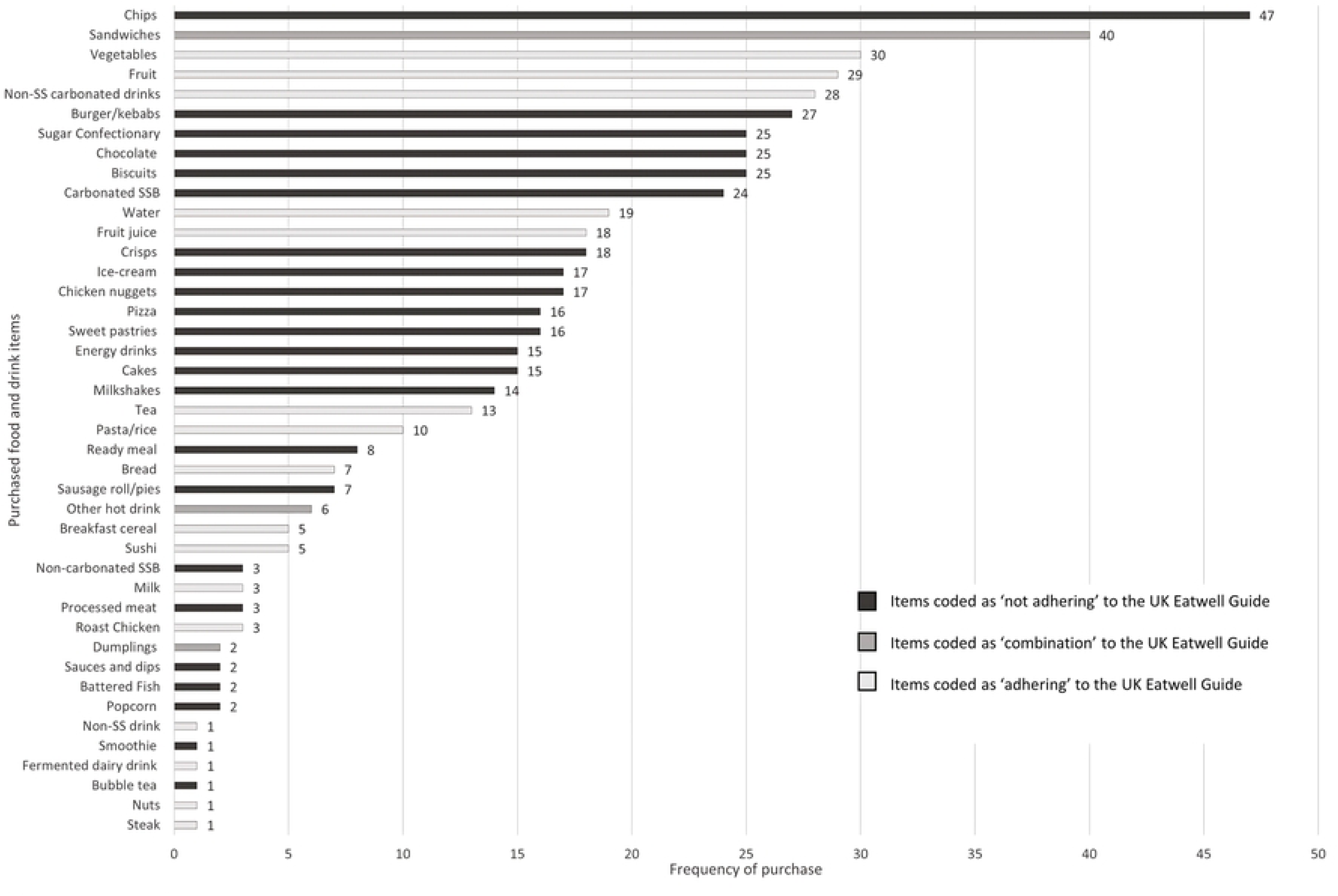
Graph illustrating the types and frequency of foods/drinks purchased by adolescents during the study.

**Fig 2.**
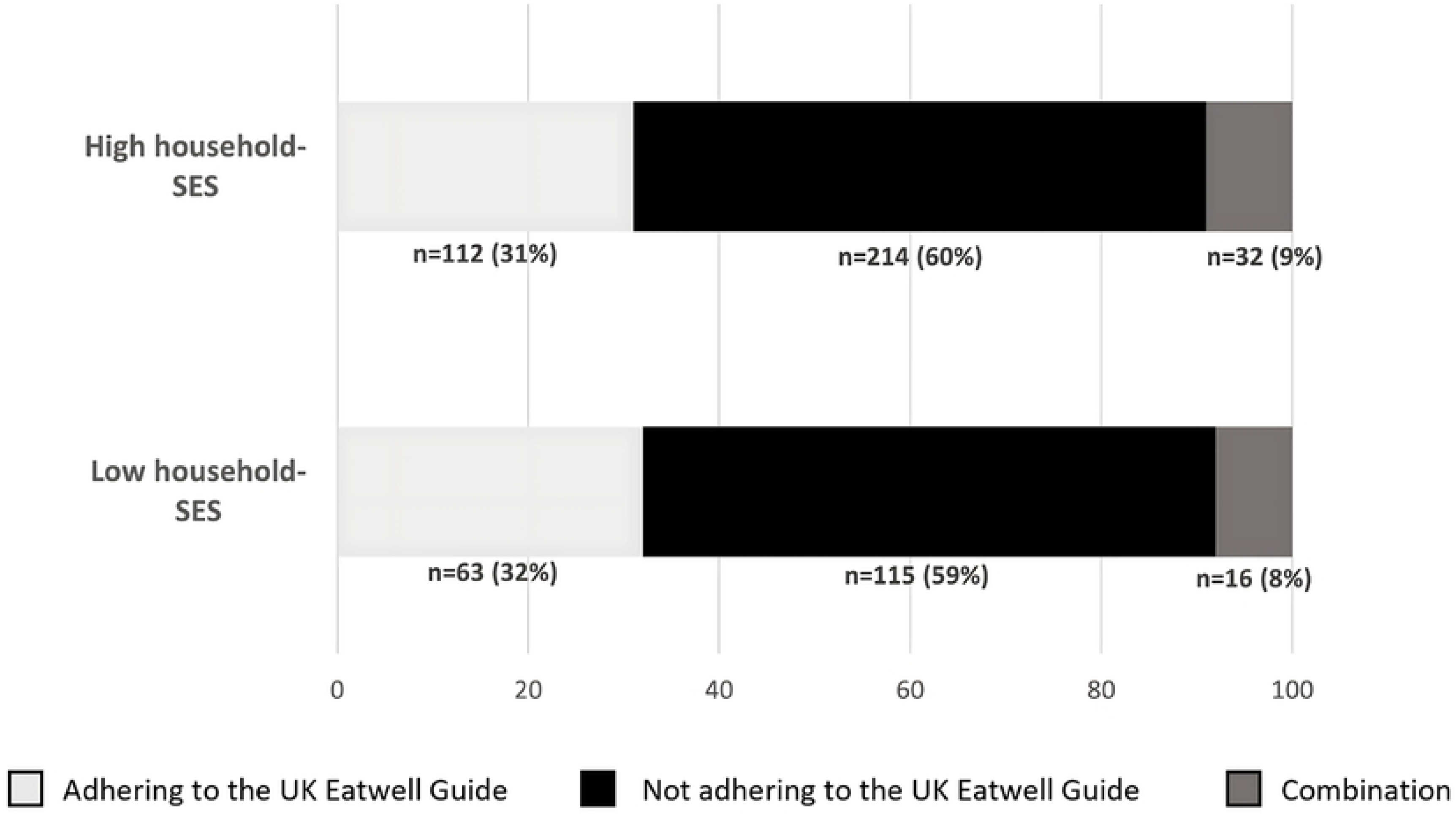
Percentages of purchases coded in UK Eatwell Guide categories by household SES.

Of the 253 food purchasing occasions, participants reported that the food and drinks purchased on 39% of these occasions were for snacks, 26% were for lunch, 26% for dinner. Only 7% were for breakfast (Table 3). Purchases bought for snacks were less healthy than those made for breakfast, lunch or dinner (p<0.001).

**Table 3.** Purchasing Healthfulness Scores on food purchasing occasions according to purpose of purchase.

|  | n | (%)* | Purchasing Healthfulness Score<sup>a</sup><br>(Median (IQR)) | p-value |
| --- | --- | --- | --- | --- |
| **Purpose of purchase** |  |  |  |  |
| Breakfast | 18 | (7) | 0.00 (-0.33, 0.50) | <0.001<sup>b</sup> |
| Lunch | 66 | (26) | 0.00 (-0.67, 0.33) |  |
| Dinner | 66 | (26) | -0.25 (-1.0, 0.00) |  |
| Snack | 98 | (39) | -1.00, (-1.0, 0.00) |  |
| Missing | 5 | (2) | - |  |

\* % of food purchasing occasions (n total = 253)
<sup>a</sup> Purchasing healthfulness score on food purchasing occasions
<sup>b</sup> Kruskal-Wallis test with adjustment for ties

The median Weekly Purchasing Healthfulness Score, reflecting the overall healthfulness of all purchases made over the study week, was −0.33 healthfulness units (IQR −0.65, 0.00) indicating that for every healthy purchase made, on average two unhealthy purchases were made. Table 4 presents the median number of weekly purchases and median Weekly Purchasing Healthfulness Score according to participants’ demographic characteristics. On average, older adolescents and girls purchased more food and drink items over the study week compared to younger adolescents (p=0.07) and boys, respectively (p=0.003). Adolescents of white ethnicity tended to purchase more items than those of other ethnicities (p=0.22). Participants from more deprived backgrounds purchased more food and drink items than those from more affluent backgrounds, but findings did not reach statistical significance (IMD p=0.20, FAS p=0.71). No notable differences were observed in Weekly Purchasing Healthfulness Score according to age, gender, ethnicity, and SES measures.

**Table 4.**
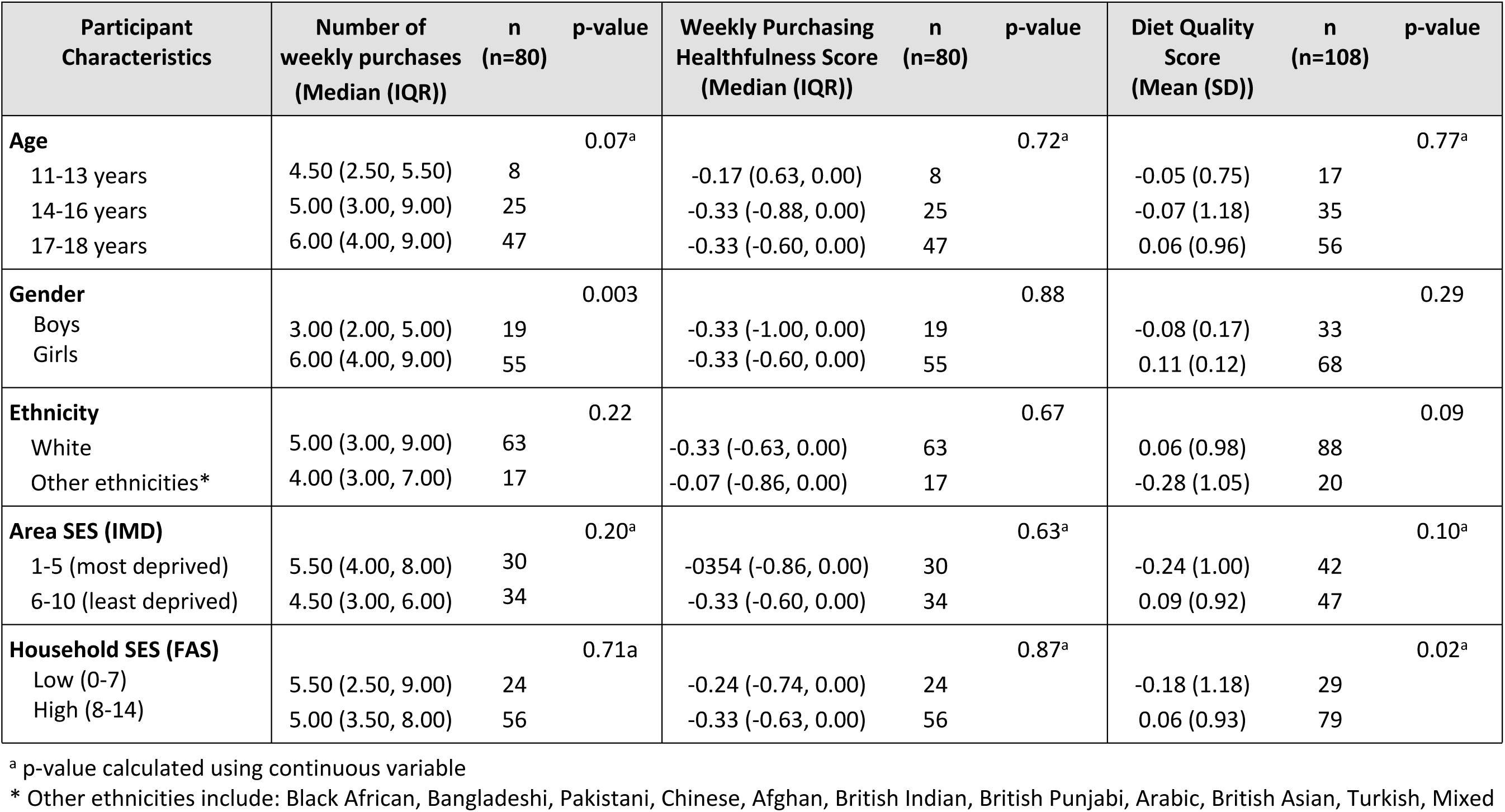
Participant purchases and diet quality scores according to characteristics.

| Participant Characteristics | Number of weekly purchases (Median (IQR)) | n (n=80) | p-value | Weekly Purchasing Healthfulness Score (Median (IQR)) | n (n=80) | p-value | Diet Quality Score (Mean (SD)) | n (n=108) | p-value |
| --- | --- | --- | --- | --- | --- | --- | --- | --- | --- |
| **Age** |  |  | 0.07<sup>a</sup> |  |  | 0.72<sup>a</sup> |  |  | 0.77<sup>a</sup> |
| 11-13 years | 4.50 (2.50, 5.50) | 8 |  | -0.17 (0.63, 0.00) | 8 |  | -0.05 (0.75) | 17 |  |
| 14-16 years | 5.00 (3.00, 9.00) | 25 |  | -0.33 (-0.88, 0.00) | 25 |  | -0.07 (1.18) | 35 |  |
| 17-18 years | 6.00 (4.00, 9.00) | 47 |  | -0.33 (-0.60, 0.00) | 47 |  | 0.06 (0.96) | 56 |  |
| **Gender** |  |  | 0.003 |  |  | 0.88 |  |  | 0.29 |
| Boys | 3.00 (2.00, 5.00) | 19 |  | -0.33 (-1.00, 0.00) | 19 |  | -0.08 (0.17) | 33 |  |
| Girls | 6.00 (4.00, 9.00) | 55 |  | -0.33 (-0.60, 0.00) | 55 |  | 0.11 (0.12) | 68 |  |
| **Ethnicity** |  |  | 0.22 |  |  | 0.67 |  |  | 0.09 |
| White | 5.00 (3.00, 9.00) | 63 |  | -0.33 (-0.63, 0.00) | 63 |  | 0.06 (0.98) | 88 |  |
| Other ethnicities* | 4.00 (3.00, 7.00) | 17 |  | -0.07 (-0.86, 0.00) | 17 |  | -0.28 (1.05) | 20 |  |
| **Area SES (IMD)** |  |  | 0.20<sup>a</sup> |  |  | 0.63<sup>a</sup> |  |  | 0.10<sup>a</sup> |
| 1-5 (most deprived) | 5.50 (4.00, 8.00) | 30 |  | -0.354 (-0.86, 0.00) | 30 |  | -0.24 (1.00) | 42 |  |
| 6-10 (least deprived) | 4.50 (3.00, 6.00) | 34 |  | -0.33 (-0.60, 0.00) | 34 |  | 0.09 (0.92) | 47 |  |
| **Household SES (FAS)** |  |  | 0.71<sup>a</sup> |  |  | 0.87<sup>a</sup> |  |  | 0.02<sup>a</sup> |
| Low (0-7) | 5.50 (2.50, 9.00) | 24 |  | -0.24 (-0.74, 0.00) | 24 |  | -0.18 (1.18) | 29 |  |
| High (8-14) | 5.00 (3.50, 8.00) | 56 |  | -0.33 (-0.63, 0.00) | 56 |  | 0.06 (0.93) | 79 |  |

<sup>a</sup> p-value calculated using continuous variable
\* Other ethnicities include: Black African, Bangladeshi, Pakistani, Chinese, Afghan, British Indian, British Punjabi, Arabic, British Asian, Turkish, Mixed

### Diet quality

Table 4 shows mean Diet Quality Scores according to demographic characteristics. Diet Quality Scores were similar across the three age groups but tended to be higher among girls compared to boys (p=0.29) and higher among adolescents of white ethnicity compared to other ethnicities (p=0.09). Diet Quality Scores were also higher among adolescents who were more affluent according to both area level SES (IMD) (p=0.10) and household SES (Family Affluence Score) (p=0.02).

#### Aim 1: To assess the associations between food purchasing behaviours and diet quality in adolescents

Table 5 presents the results of linear regression analyses, testing the association between food purchasing and diet quality. Unadjusted models showed no relationship between number of weekly food purchases and diet quality, and this relationship remained when models were adjusted for age, gender, ethnicity, and household SES. When the healthfulness of these purchases was considered, a positive association with diet quality was observed. Higher Weekly Purchasing Healthfulness Scores were associated with higher Diet Quality Scores (β 0.48 SDs/healthfulness unit, (95%CI 0.01, 0.96) p=0.05). These results were attenuated after adjustment (β 0.36 SDs/healthfulness unit, (95%CI −0.15, 0.87) p=0.16).

**Table 5.** Linear regression results for association between food purchasing (exposure) and diet quality (SDs) (outcome)

| Exposure variable | Unadjusted $\beta$<br>(95% CI) | p-value | Adjusted $\beta$<br>(95% CI)* | p-value |
| --- | --- | --- | --- | --- |
| Number of weekly purchases | 0.00<br>(-0.04, 0.04) | 0.98 | 0.01<br>(-0.04, 0.04) | 0.78 |
| Weekly Purchasing Healthfulness Score (healthfulness units) | 0.48<br>(0.01, 0.96) | 0.05 | 0.36<br>(-0.15, 0.87) | 0.16 |

\*Models adjusted for age, gender, ethnicity, and household SES.

#### Aim 2: To assess whether SES moderates the relationship between food purchasing behaviours and diet quality

Adding the interaction term to the regression models showed that the association between Weekly Purchasing Healthfulness Score and diet quality was stronger among participants with low household SES compared to those with high household SES (p for interaction=0.01) (Fig 3). Adjusted stratified analyses showed a positive relationship between the healthfulness of weekly purchases and diet quality for adolescents from households of lower SES (β 1.04 SDs/healthfulness unit, (95%CI 0.26, 1.81) p=0.01). Among adolescents with higher household SES, no clear association was observed (β 0.27 SDs/healthfulness unit, (95%CI −0.25, 0.81) p=0.31).

**Fig 3.**
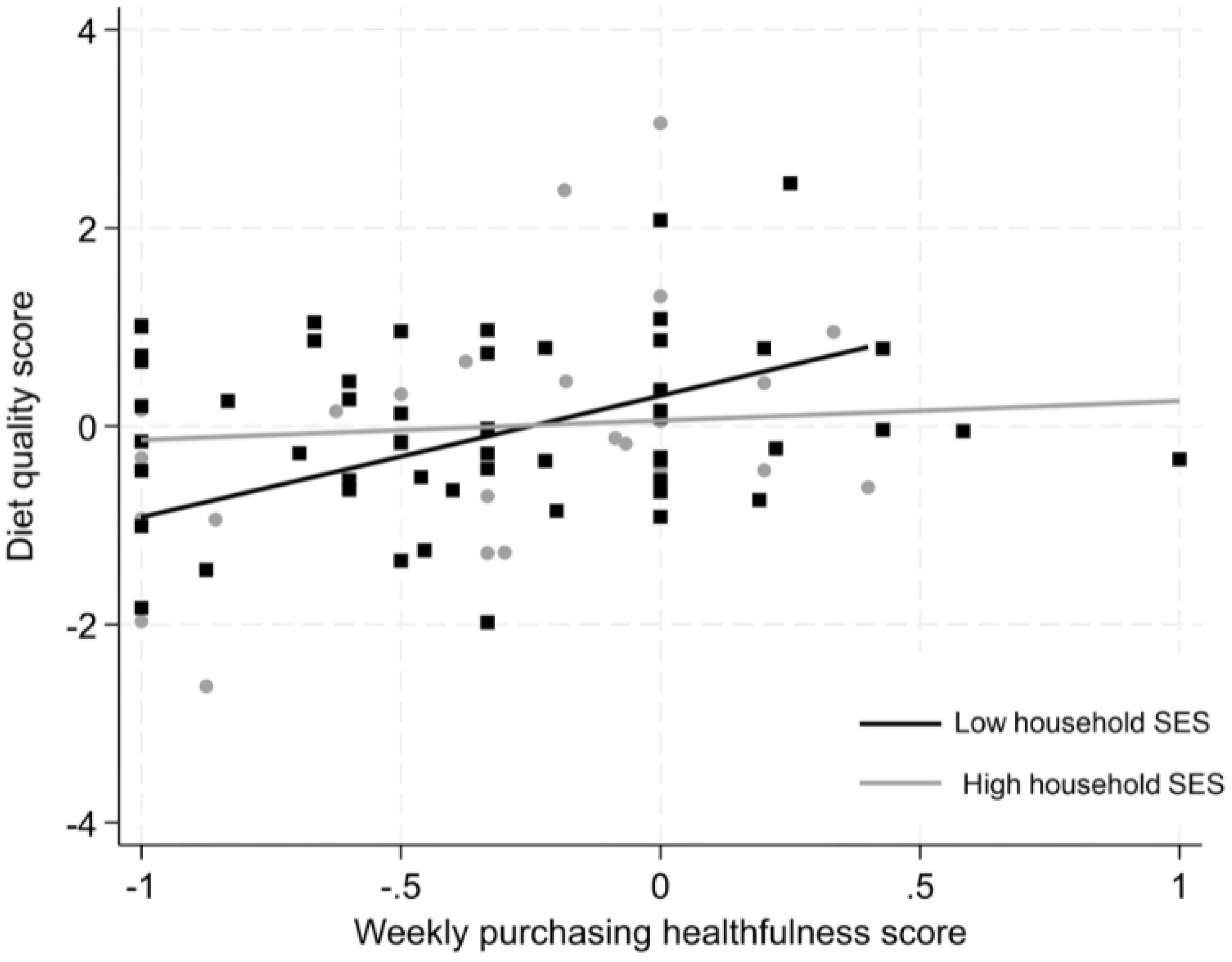
Diet quality scores by weekly purchasing healthfulness score according to household SES.

## Discussion

### Summary of key findings

This study provides novel insight into the independent food purchasing behaviours of adolescents and their relationship with overall dietary quality. The majority of food purchases made by adolescents participating in this study, when they were away from home and school, were not aligned with national healthy eating guidelines and had the potential to negatively impact their health. Adolescents in this study reported that 39% of their food and drink purchases were snacks rather than a main meal and these snack purchases were less healthful products according to the UK Eatwell Guide. Adolescents who made more healthful purchases over the study week tended to have better quality diets and this was particularly true for adolescents experiencing socio-economic disadvantage.

### Interpretation and comparison with previous research

Previous research has shown that adolescents source the majority of their food from home and school (6, 7). This study demonstrates, however, the important role of independent food purchases from food outlets, outside of home and school environments, on adolescents’ overall dietary quality. This study also provides insights into the types of foods adolescents purchase for themselves. These purchasing data offer a more nuanced understanding than previous studies which have reported adolescent purchasing of only specific food items (e.g. sugar-sweetened beverages) or unspecific categories such as junk food or snack food (27, 28). While the majority (62%) of the purchases made by participants in this study were categorised as ‘not adhering’ to the UK Eatwell Guide, adolescents do purchase, albeit in smaller quantities, food and drink items which are aligned with healthy eating guidelines. Further research is needed to understand the factors that encourage adolescents to purchase these more healthful items, over less healthy alternatives, and if these factors vary by SES.

A disparity in dietary behaviours between higher and lower socioeconomic groups has been well documented in the literature. Adolescents from disadvantaged backgrounds are more likely to have poorer quality diets consisting of higher intakes of energy dense, nutrient poor foods and lower intakes of fruits and vegetables (29–31). Additionally, previous research illustrates that food purchases made by adults experiencing socio-economic disadvantage are less healthy than more affluent adults (32, 33). The findings from this study differ from those of previous studies because they show no variation in the total number or the healthfulness of purchases made by adolescents from high and low SES households. Adolescents who purchased more healthful products, however, were more likely to have better quality diets. There was no clear relationship between the number of purchases made by adolescents and their diet quality. Together these findings suggest that it is not how often adolescents purchase food for themselves that is an important contributor to their diet quality but the healthfulness of the purchases that they make regardless of how often they shop.

For adolescents experiencing disadvantage based on household SES, the food purchases they made for themselves played a stronger role in their overall diet quality compared to adolescents from more affluent families. One potential explanation for this finding is that independent food purchases make up a larger proportion of the overall diets of adolescents from more deprived backgrounds. Since these purchases tend not to conform to healthy eating guidelines, they are likely to reduce the overall quality of these young people’s diets. Another driver of this difference may be that adolescents from less affluent backgrounds are not exposed to the same health-promoting foods at home as those living in more affluent households. Previous research conducted among Scottish families with teenagers found that even though health is valued when making food choices, families from lower SES were driven to make choices that ensured all family members were fed in a quick and acceptable way (34). Backett-Milburn et al (2006) described how competing priorities within families with lower-incomes means nutrition and health fall below other factors such as affording everyday essentials, safety and worries about children engaging in other risky behaviours such as drugs, alcohol and smoking (35). Additionally, a report published in 2023 further highlighted how the cost-of-living crisis has made healthy eating seem more unattainable for many low income household due to rising food and energy costs (36). This research has also illustrated that families experiencing low-income are driven to select less healthful foods because of poor housing and food preparation facilities, having less autonomy over their working practices and to fulfil a range of social, emotional and cultural needs; financial constraints mean they are less able to fulfil these needs in other ways (36, 37). In contrast, results from a qualitative study have shown families experiencing higher SES have greater opportunity to offer experiences incorporating healthy foods and were more likely to discuss the nutritional significance of food choices in the home (34). These families described limiting their intake of sugary and fatty foods while simultaneously encouraging younger family members to try a variety of different ‘spicy’ or ‘exotic’ foods to widen their taste preferences when they became adults (34). Taken together, this body of evidence suggests that adolescents from lower SES do not have the same opportunities to consume a healthy or varied diet in the home as those from higher SES. Therefore, the purchases adolescents experiencing low SES make for themselves, when out of the home, may be particularly important targets for improving their quality of their diets.

### Research and policy implications

This study suggests that finding ways to promote and encourage more healthful independent food choices among adolescents, particularly those from disadvantaged backgrounds, is likely to be an important strategy in improving their overall dietary quality and, in turn, reducing dietary inequalities. Given the challenges faced by those from low-income households in relation to affording and acquiring heathy foods, the responsibility to support adolescents to eat more healthfully should not fall solely on parents but should be the responsibility of wider society. Future public health strategies and food policies will be most effective if they aim to support adolescents to acquire healthy food from multiple different sources, including neighbourhood and school environments.

To enhance the accessibility of healthy foods for adolescents experiencing low SES, zoning restrictions of fast-food outlets and takeaways in disadvantaged areas may be particularly important. Previous research has shown that the most deprived areas in the UK are those that have the greatest access to unhealthy food outlets (38, 39), zoning restrictions are likely to particularly benefit those living and spending time in more deprived communities. Further to this, changes to the environments inside the food outlets adolescents use most often could also help support more healthful purchases. The UK government have introduced restrictions on placing foods high in fat, sugar and salt in prominent areas of food stores. In addition, restrictions on the promotion of these items have been proposed (40). Such food policies offer the opportunity to support more healthful purchases among all young people as long as they are implemented effectively across all types of retail outlets, neighbourhoods and regions (41).

Snack purchases made up the largest proportion of purchases by adolescents in the current study and tended to be less healthful. This finding is consistent with research from the Netherlands which showed adolescents mainly purchased snack food items from supermarkets close to school and spent, on average €2.30 (∼USD 2.45) (42). Qualitative research shows that adolescents favour quick, single-serve, low-cost snack options (43). Future research and food policy development would benefit from targeting the availability of healthier snack options that are affordable and appealing to adolescents. These healthier snack options should be available in the food outlets adolescents regularly visit and offer a possibility for new product development.

While not specifically measured in the current study, improving the accessibility and desirability of healthy free school meals is recognised as an important strategy to support young people to source and eat more healthful foods. Currently, in England, eligibility for free school meals is means-tested but the low-income threshold means that many young people who are from working families but still living in poverty are not eligible for the scheme. Widening the eligibility criteria as well as ensuring free school meals meet the school food standards could help reduce dietary inequalities, particularly if implemented concurrently to improvements to neighbourhood food environments (44). By encouraging better food consumption within schools, through enhanced provision of free school meals, the purchases of unhealthy snacks or meals after school may also reduce.

### Strengths and limitations

This study is novel in its assessment of adolescents’ independent food purchasing behaviours in the community setting. The EMA techniques used to collect purchasing data are a strength of the study because they allowed participants to record data as close as possible to the behaviour of interest, helping to reduce the risk of recall bias. It was not possible in this study to determine the completeness of the purchasing data because of the uncertainty regarding those who made no recordings. It was not possible to determine whether they were not using the app or did not make any purchases. While the purchasing data provided a more detailed description of purchases made by adolescents than many published studies, data about food brands, portion sizes and nutritional information were not collected. It is therefore possible that purchases could have been miscategorised in terms of healthfulness.

The study’s small sample size reduces statistical power and the use of a convenience sample limits generalisability of the findings. The direction of the study findings, however, show promise and justify replication in a larger and more diverse population.

## Conclusion

This study demonstrated that adolescents predominately purchase food and drink items that are not aligned with healthy eating guidelines. Such purchasing decisions are more important for overall dietary quality among adolescents experiencing disadvantage. Findings from this study suggest that introducing food policies that support adolescents to make healthy food purchases from multiple different food environments (namely community and school) could act to reduce inequalities and improve health and well-being among the next generation of adults. Such policies may include zoning policies to limit the number of unhealthy food outlets in deprived areas, addressing placement within shops of unhealthy food promotions, widening access to free school meals and developing healthy, affordable and socially desirable snack options.

## Data Availability

Due to ethical restrictions imposed in the interest of protecting participant confidentiality, the data underlying this study are available upon request. Researchers wishing to use the data can make apply to the research team by emailing. Subject to approval that the intended purpose is compatible with the study’s ethical approval and formal agreements regarding confidentiality and secure data storage being signed, the data would then be provided.

## Acknowledgements

The authors would like to thank the teachers and youth leader who helped with the recruitment process for this study as well as all the young people who participated. The authors would also like to thank Patsy Coakley for her assistance in preparing the data for this study.

## Author Contributions

S. Shaw: Conceptualization, Data curation, Formal Analysis, Funding acquisition, Investigation, Methodology, Project administration, Writing-original draft, Writing-reviewing and editing. S. Crozier: Formal Analysis, Supervision, Writing-reviewing and editing. C. Cooper: Supervision, Writing-reviewing and editing. D. Smith: Supervision, Writing-review & editing. M. Barker: Conceptualization, Funding acquisition, Supervision, Writing-review & editing. C.Vogel: Conceptualization, Funding acquisition, Methodology, Supervision, Writing-review & editing.

